# Diffuse white matter abnormality in very preterm infants reflects reduced brain network efficiency

**DOI:** 10.1101/2020.06.27.20141713

**Authors:** Julia E. Kline, Venkata Sita Priyanka Illapani, Hailong Li, Lili He, Nehal A. Parikh

**Affiliations:** Perinatal Institute, Cincinnati Children’s Hospital Medical Center, Cincinnati, OH; Department of Pediatrics, University of Cincinnati College of Medicine, Cincinnati, OH; Center for Perinatal Research, The Research Institute at Nationwide Children’s Hospital, Columbus, OH

## Abstract

Between 50-80% of very preterm infants (≤32 weeks gestational age) exhibit increased white matter signal intensity on T2 MRI at term-equivalent age, known as diffuse white matter abnormality (DWMA). A few studies have linked DWMA with microstructural abnormalities, but the exact relationship remains poorly understood. We used graph theory methods to relate DWMA extent to measures of efficient information processing at term in a representative cohort of 343 very preterm infants. We performed anatomic and diffusion MRI at term and quantified DWMA volume using our novel, semi-automated algorithm. From structural connectomes, we calculated graph theory metrics: local efficiency and clustering coefficient, which measure the ability of groups of nodes to perform specialized processing, and global efficiency, which assesses the ability of brain regions to efficiently combine information. We computed partial correlations between these measures and DWMA volume, adjusted for confounders. Increasing DWMA volume was associated with decreased global efficiency of the entire brain network (r= - 0.27, p= 8.36E-07) and decreased local efficiency and clustering coefficient within individual networks supporting cognitive, linguistic, and motor functions. We show that DWMA is associated with widespread decreased brain network connectivity in very preterm infants, suggesting it is pathologic and likely has adverse developmental consequences.

## Introduction

Up to 80% of very preterm infants (gestational age (GA) ≤ 32 weeks) exhibit increased white matter signal intensity on T2-weighted MRI at term-equivalent age ^1,2^. This hyperintensity, which has been referred to as diffuse excessive high signal intensity (DEHSI), and more recently and descriptively as diffuse white matter abnormality (DWMA), has been investigated in relation to neurodevelopmental outcomes in premature infants with mixed results. Some studies have shown no significant relationship between DWMA and later neurodevelopmental deficits, including cognitive, language, and motor impairments ^2–10^. Conversely, other studies ^11–14^, especially those that quantified DWMA extent at term ^15,16^, have identified significant associations with neurodevelopmental outcomes, particularly cognition and language.

The underlying cellular pathology and brain connectivity alterations in DWMA have not been fully delineated. However, abnormal white matter microstructure from diffusion MRI ^1,17^ and aberrant functional connectivity from resting state functional MRI ^18^ have been documented at term in pilot studies of preterm infants with DWMA. A small postmortem histopathology case series ^19^ reported far fewer oligodendroglial cells and axons in the brains of very preterm infants in DWMA regions compared to the brains of control infants. Based on these results, we postulated that DWMA is associated with widespread brain network disorganization in very preterm infants.

Graph theory is a powerful mathematical method that can query the organizational structure and information processing capacity of complex networks like the human brain ^20–24^. The brain’s connectome can be modeled as a set of regions (nodes) and their pair-wise associations (edge weights), which may represent physical connections or effective or functional connectivity values. From such connectivity matrices, several metrics can be calculated that quantify the ease of information transfer and processing within the network. A subset of these: global efficiency (E_glob_), local efficiency (E_loc_), and clustering coefficient (CC), have been shown to be altered by preterm birth ^25–27^, and these alterations may be the antecedents of lifelong neurodevelopmental impairment ^28^. For an in-depth discussion of graph theory and its implications for network neuroscience, refer to the work of Rubinov and Sporns ^21^.

Our main objective was to quantify DWMA in a regional, population-based cohort of very preterm infants using our published semi-automated algorithm and to examine the relationship between DWMA volume and graph theory metrics of efficient information processing: E_glob_, E_loc_, and CC. We hypothesized that 1) all three metrics would be adversely influenced by several key risk factors of preterm brain development and 2) graph theory metrics of efficiency would decrease with increasing extent of objectively-quantified DWMA, particularly in nodes related to cognition, language, and motor ability.

## Methods

### Subjects

We enrolled a multicenter, prospective cohort of 343 very preterm infants from five level-III Greater Cincinnati area neonatal intensive care units: 1) Cincinnati Children’s Hospital Medical Center (CCHMC), Cincinnati’s primary academic pediatric referral service for the sickest neonates; 2) University of Cincinnati Medical Center (UCMC), Cincinnati’s primary academic hospital for high-risk maternal referral; 3) Good Samaritan Hospital (GSH); 4) Kettering Medical Center (KMC); and 5) St. Elizabeth’s Healthcare (SEH); the latter three represent non-academic sites. Subjects were recruited between June 2017 and October 2019 and were excluded if they had cyanotic heart disease or chromosomal or congenital anomalies affecting their central nervous system. Infants who were hospitalized and mechanically ventilated on more than 50% supplemental oxygen at 45-weeks postmenstrual age (PMA) were also excluded. The Cincinnati Children’s Hospital Institutional Review Board approved this study, resulting in approval at the other sites due to reciprocity agreements. A parent or guardian of each infant gave written informed consent before enrollment.

### MRI data acquisition

All study infants were imaged during unsedated sleep between 40- and 44-weeks postmenstrual age on a 3T Philips Ingenia scanner with a 32-channel receiver head coil. A skilled neonatal nurse and neonatologist were both present for any scans requiring positive pressure airway support. Each infant was fed 30 minutes prior to MRI, fitted with silicone earplugs to mitigate scanner noise (Instaputty, E.A.R. Inc, Boulder, CO), and swaddled in a blanket and a vacuum immobilization device (MedVac, CFI Medical Solutions, Fenton, MI) to promote natural sleep. We acquired MRI data as follows: diffusion MRI: echo time 88 ms, repetition time 6972 ms, flip angle 90°, field of view 160 × 160 mm^2^; 80 × 79 matrix; and 2-mm contiguous slices; scan time 5:58 min. 36 directions of diffusion gradients were applied with a b value of 800 s/mm^2^; low b-value = 0 (4 b0 images were acquired with posterior-anterior phase encoding and 1 b0 image was acquired with anterior-posterior phase encoding); axial T2-weighted image: echo time 166 ms, repetition time 18567 ms, flip angle 90°, voxel dimensions 1.0 × 1.0 × 1.0 mm^3^, scan time 3:43 min; 3-dimensional magnetization-prepared rapid gradient echo: echo time 3.4 ms, repetition time 7.3 ms, flip angle 11°, voxel dimensions 1.0 × 1.0 × 1.0 mm^3^, scan time 2:47 min; sagittal SWI: echo time 7.2 ms, repetition time 29 ms, flip angle 17°, voxel dimensions 0.57 × 0.57 × 1.0 mm^3^, scan time 3:27 min.

### DWMA quantification

We quantified whole-brain DWMA on T2 images using our previously-described semi-automated software ^29^. Briefly, after preprocessing the T2 images by bias field correction and intensity normalization and performing tissue segmentation via a unified algorithm ^30^ with the guide of a neonatal atlas ^31^, our program designated white matter voxels as DWMA if their intensity was greater than 1.8 standard deviations above the mean intensity for all grey and white matter voxels, a cutoff determined by our previous research ^16^ (Fig. 1). For each subject, we normalized whole-brain DWMA volume by the infant’s total white matter volume, to correct for the effect of differing head sizes.

**Figure 1:**
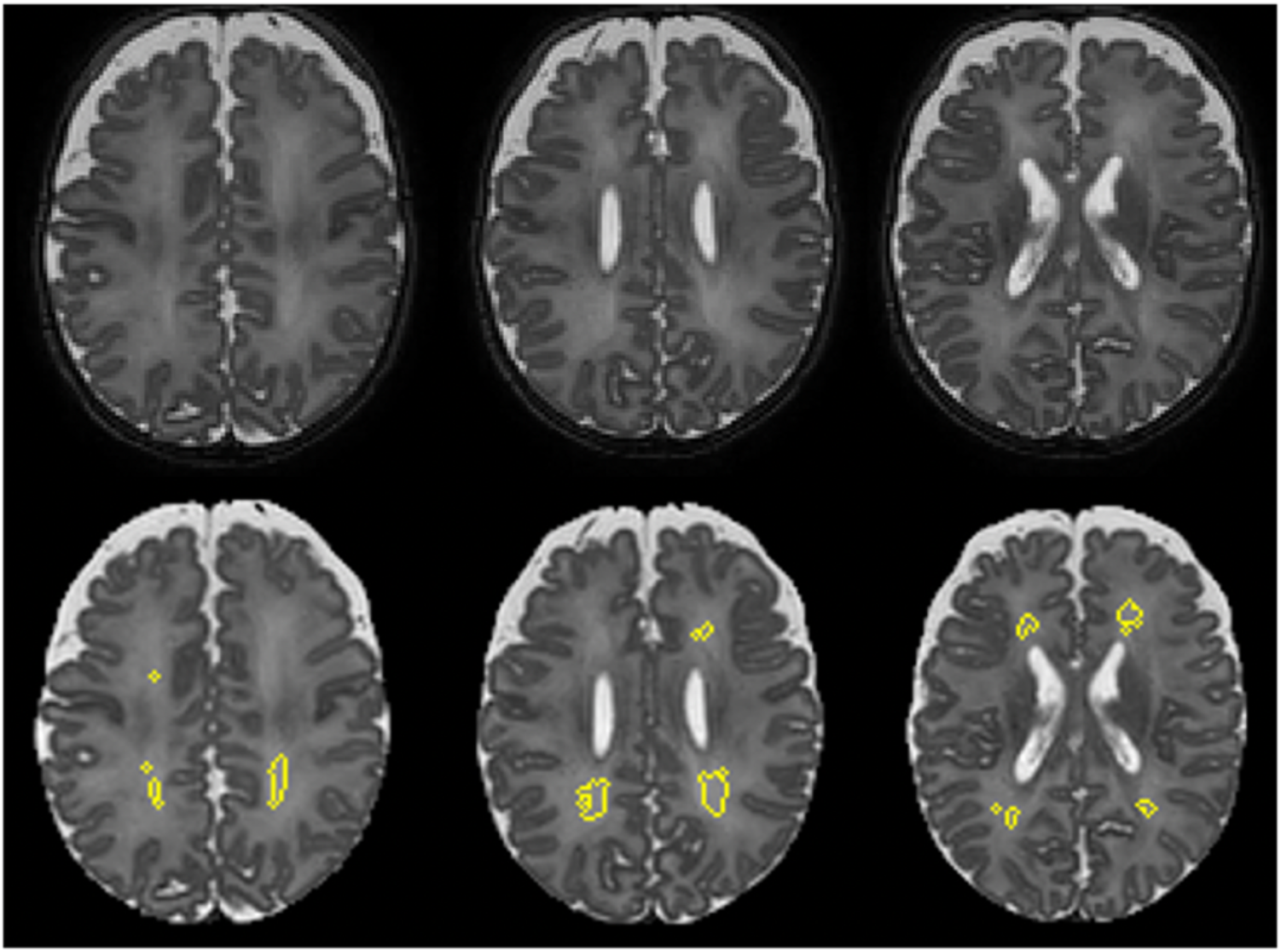
Example Semi-automated DWMA Segmentation. The top row shows three slices from a T2 MRI image of the same male infant; gestational age 25.4 weeks, postmenstrual age at MRI 41.7 weeks. The bottom row shows the same slices with clusters of voxels designated as DWMA by our algorithm circled in yellow.

### Brain abnormality scoring

We used the standardized scoring system developed by Kidokoro ^32,33^ to derive a brain abnormality score for each study subject. The overall score is the sum of four individual scores quantifying the extent of abnormalities in 1) the cortical grey matter, 2) the cerebral white matter, 3) the deep grey matter, and 4) the cerebellum. A single pediatric neuroradiologist (B.K.), who was masked to the clinical history of the study subjects, performed all qualitative and quantitative MR image assessments.

### Diffusion MRI processing

We first rejected any diffusion-weighted scan for which more than four volumes exhibited motion artifact-induced displacement greater than the width of one voxel. For the remaining scans, we used standard FSL (http://fsl.fmrib.ox.ac.uk/fsl/fslwiki/) routines to preprocess the diffusion data. We corrected for susceptibility-induced distortions using the reverse phase-encoded b0 image, and we also corrected for movement and eddy current-induced distortions. The Diffusion Toolkit software by Trackvis (www.trackvis.org) generated a tensor model in each brain voxel and produced fractional anisotropy (FA) maps in diffusion space. (FA, one of the most commonly-derived metrics from the diffusion tensor model, measures the degree of directionality of the white matter fibers and can be thought of as a surrogate marker of white matter integrity.) From the FA maps, we performed whole-brain, deterministic fiber tracking. We linearly aligned each subject’s T2 image to their diffusion b0 image, to produce an image with more clearly-delineated anatomical boundaries than the original b0. We then aligned the 90-region automated anatomical labeling (AAL) infant template ^31,34^ to this new image, to create parcellated brain maps in diffusion space.

### Brain structural connectome construction

We used MRTRIX software (http://www.mrtrix.org) to create connectivity matrices by first extracting the FA value at each discrete point along each white matter streamline and then generating symmetrical 90x90 matrices, with the on-diagonal elements representing the mean FA for all tracts traversing each region and the remaining elements representing the mean FA for all tracts connecting each pair of regions. These values are thus the edge weights in weighted, undirected connectivity matrices. To avoid spurious associations based on noise in the diffusion image and because many regional pairs are not biologically plausible, we reassigned matrix elements with mean FA < 0.1 to zero.

### Graph theory metrics

We used the open-source, MATLAB-compatible Brain Connectivity Toolbox (www.brain-connectivity-toolbox.net) to calculate the graph theory metrics of interest from connectivity matrices. G*lobal efficiency* (E_glob_) of a network is defined as its average inverse shortest path length ^35^. It is a measure of functional integration, or the ability of distant regions to rapidly combine specialized information. *Local efficiency* (E_loc_), on the other hand, is a measure of functional segregation, or the ability of discrete groups of nodes to carry out specialized processing. As described by Latora and Marchiori ^36^, E_loc_ quantifies the degree of connectedness between a node’s neighbors. The *clustering coefficient* (CC) of a node is the fraction of the node’s neighbors that are neighbors of each other. CC measures the ability of densely interconnected groups of nodes to engage is specialized information processing, and therefore is also a measure of functional segregation. We computed global efficiency for each connectome and local efficiency and clustering coefficient for each brain node

### Statistical analysis

For our major analysis, in Stata 16.0 (StataCorp, College Station, TX), we computed partial correlations and the associated p-values for DWMA volume and graph theory measures (E_glob_, E_loc_, and CC), with and without the effects of PMA at MRI, gestational age, sex, structural brain abnormality ^32^, and birth hospital removed. We applied Benjamini-Hochberg false discovery rate (FDR) correction to account for all observations made (181 graph theory metrics tested), with an accepted FDR of 5%. The investigator who performed the DWMA quantification was blinded to the results of the graph theory analysis, and vice versa. The brain networks were visualized using BrainNet Viewer (http://www.nitrc.org/projects/bnv/).

We also performed two secondary analyses, to confirm the validity of our graph theory metrics and to assess the effect of attrition on our cohort. First, to determine if graph theory measures of had the expected relationship with known risk factors, we examined the correlations between global efficiency, mean local efficiency, and mean clustering coefficient and two major risk factors of prematurity: brain abnormality score and gestational age. Second, to identify possible exclusion bias, we compared the baseline characteristics of our final cohort to the excluded infants. We used Fischer’s exact test for binary variables and either Student’s t-test or a Mann-Whitney U test for continuous variables, depending on normality. All applicable significance tests were two-tailed, and a p-value of <0.05 indicated significance.

## Results

Out of an initial cohort of 343 very preterm infants, eight were excluded due to suboptimal alignment with the AAL infant template ^31,34^, resulting from artifacts on diffusion-weighted MRI (n=7) or deformational scaphocephaly (n=1). An additional 11 subjects were excluded due to ventriculomegaly; in extreme cases very little brain tissue was spared for DWMA detection. Our final cohort comprised 324 infants with high-quality diffusion data and objectively-quantified DWMA. The mean (SD) postmenstrual age at MRI scan was 42.8 (1.3) weeks. Compared to our final cohort, excluded infants were more likely to have moderate or severe brain injury, which was expected due to the nature of the exclusions. All other baseline characteristics were comparable between the two groups. Figure 1 shows an example DWMA segmentation for a subject in the final cohort. Table 1 summarizes the clinical characteristics of both cohorts as well as their statistical differences.

**Table 1.** Baseline characteristics of the final very preterm cohort and the excluded infants.

| Characteristics | Final Cohort<br>(n=324) | Excluded<br>(n=19) | p |
| --- | --- | --- | --- |
| Antenatal steroids (completed course), n (%) | 299 (92.3) | 18 (94.7) | 1.00 |
| Gestational age, weeks, mean (SD) | 29.3 (2.5) | 28.4 (2.1) | 0.09 |
| Birth weight, grams, mean (SD) | 1308.6 (459.4) | 1174.1(311.5) | 0.10 |
| Male, n (%) | 162 (50.0) | 5 (26.3) | 0.06 |
| Severe retinopathy of prematurity*, n (%) | 18 (5.6) | 1 (5.3) | 1.00 |
| Bronchopulmonary dysplasia, n (%) | 115 (35.5) | 11 (57.9) | 0.08 |
| Late onset sepsis, n (%) | 30 (9.3) | 3 (15.8) | 0.41 |
| Postnatal steroids (dexamethasone), n (%) | 32 (9.9) | 3 (15.8) | 0.43 |
| Maternal education (college degree or higher), n (%) | 154 (47.5) | 9 (47.4) | 1.00 |
| Private insurance, n (%)** | 155 (48.3) | 5 (26.3) | 0.10 |
| Household income above \$60,000, n (%) | 159 (49.1) | 9 (47.4) | 1.00 |
| Moderate to severe injury on structural MRI, n (%) | 30 (9.3) | 9 (47.4) | <0.001 |

\* 1 included subject did not have retinopathy of prematurity data.
\*\* 3 included subjects did not have insurance data.

To illustrate the overall structure of our preterm cohort’s brain network, Figure 2 displays the mean connectome/connectivity matrix of the 324 included infants. Although we computed our graph theory metrics using a threshold of FA≥0.1, in this figure we display only the strongest subset of connections (those with mean FA≥0.2), as rendered by BrainNet Viewer (http://www.nitrc.org/projects/bnv/).

**Figure 2:**
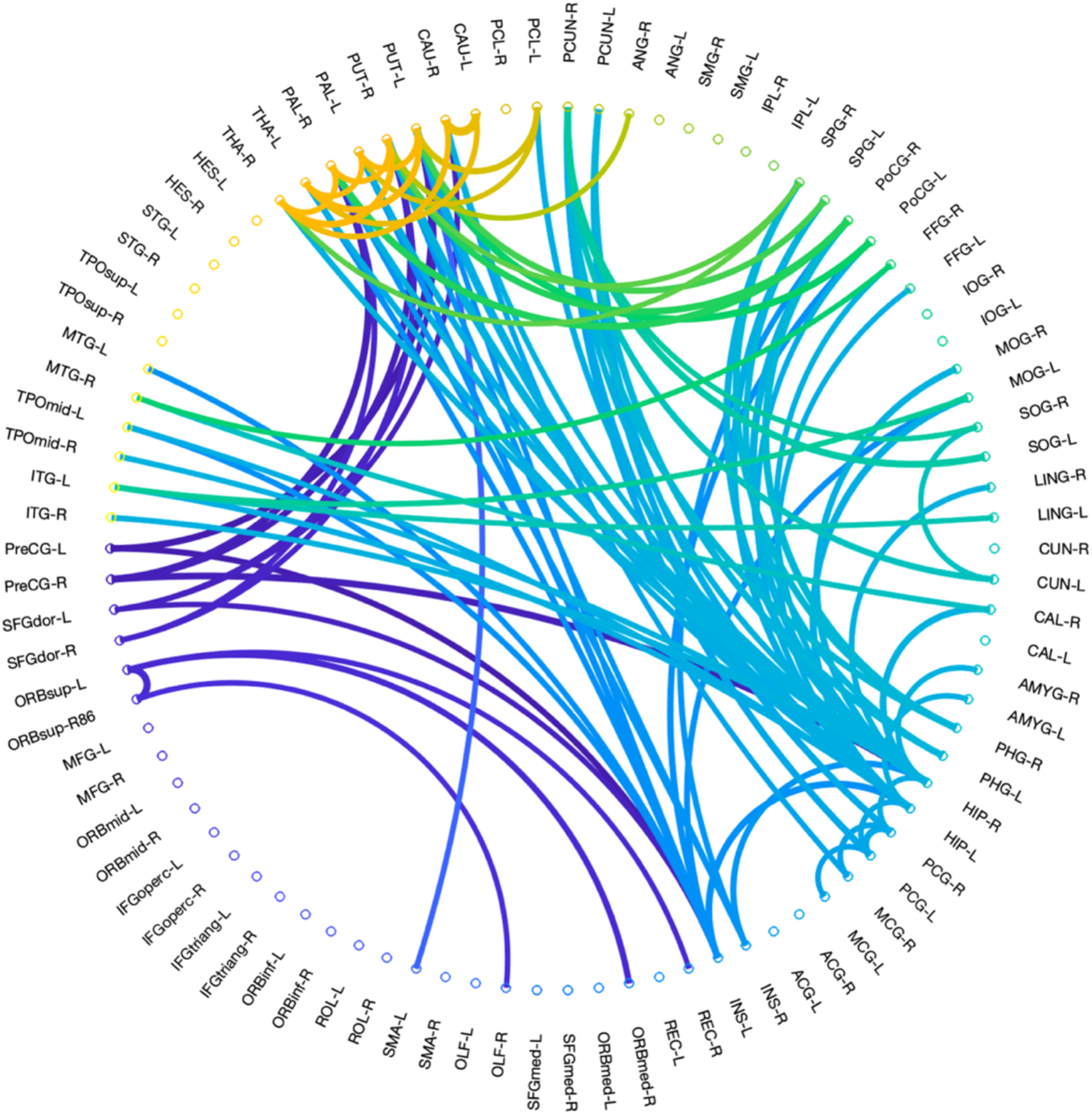
Mean Connectome of Very Preterm Cohort. The mean connectome for the final 324 subjects displayed as a circular graph composed of 90 nodes. Only connections with mean FA≥0.2 are displayed here, for easy visualization of the strongest subset of connections within the very preterm brain network. The color of the connections is arbitrary.

As expected, mean graph theory measures were significantly influenced by major risk factors of prematurity. For our final cohort, the median brain abnormality score was 2, with an interquartile range of 5. 66.7% of the cohort had no brain abnormality (scores of 0-3), 22.2% had mild abnormality (scores of 4-7), 5.9% had moderate abnormality (scores of 8-11), and 5.3% had severe abnormality (scores ≥12). Brain abnormality score was negatively correlated with global efficiency (r= −0.36, p= 9.43E-12), mean local efficiency (r= −0.30, p=2.63E-08) (Supplementary Fig. S1), and mean clustering coefficient (r= −0.25, p= 4.02E-06) (Supplementary Fig. S1). Gestational age at birth was positively correlated with global efficiency (r= 0.16, p= 0.004) but not with mean local efficiency or mean clustering coefficient.

As hypothesized, in our cohort of very preterm infants, global efficiency of the entire brain network was negatively correlated with DWMA volume (r= −0.27, p= 8.36E-07). The relationship remained robust after correcting for gestational age, brain abnormality score, sex, PMA at MRI, and birth hospital (p=2.73E-03). Mean local efficiency of the entire brain network was negatively correlated with DWMA volume (r= −0.33, p=6.64E-10), and this relationship remained significant after covariate adjustment (p= 3.33E-05). Regionally, local efficiency was negatively correlated with DWMA throughout many regions of the very preterm brain (Fig. 3, Table 2), taking into account covariate adjustment and FDR correction. Likewise, mean clustering coefficient was negatively correlated with DWMA volume in a univariate fashion (r= - 0.36, p=3.42E-11) and after adjustment for covariates (p=3.26E-6). Regionally, CC was negatively correlated with DWMA in several nodes, after covariate adjustment and FDR correction (Fig. 3, Table 2). There were no significant positive associations between DWMA volume and any graph theory metric tested.

**Table 2.** Nodes in which DWMA volume was significantly associated with local efficiency (LE) or clustering coefficient (CC), corrected for false discovery rate. NS means non-significant.

| Network Node | LE (p value) | CC (p value) | Network Membership |
| --- | --- | --- | --- |
| Precentral gyrus left **PreCG-L** | 1.65E-02 | 1.21E-02 | Motor |
| Superior frontal gyrus (dorsal) left **SFGdor-L** | 8.39E-03 | NS | Cognition |
| Superior frontal gyrus (dorsal) right **SFGdor-R** | NS | 5.83E-03 | Cognition |
| Orbitofrontal cortex (superior) left **ORBsup-L** | 8.43E-04 | NS | Cognition |
| Orbitofrontal cortex (superior) right **ORBsup-R** | 7.42E-05 | NS | Cognition |
| Middle frontal gyrus left **MFG-L** | 4.44E-04 | 1.38E-03 | Cognition |
| Orbitofrontal cortex (middle) left **ORBmid-L** | 1.41E-04 | NS | Cognition |
| Orbitofrontal cortex (middle) right **ORBmid-R** | 1.17E-03 | 5.26E-03 | Cognition |
| Inferior frontal gyrus (opercular) left **IFGoperc-L** | 2.84E-05 | 1.27E-04 | Language |
| Inferior frontal gyrus (triangular) left **IFGtriang-L** | 5.88E-03 | NS | Language |
| Orbitofrontal cortex (inferior) right **ORBinf-R** | 7.57E-03 | NS | Cognition |
| Rolandic operculum left **ROL-L** | 7.49E-07 | 3.15E-03 | Language, Motor |
| Rolandic operculum right **ROL-R** | 9.01E-03 | NS | Language, Motor |
| Olfactory left **OLF-L** | 1.31E-02 | NS |  |
| Insula left **INS-L** | 5.56E-07 | 5.16E-03 | Cognition |
| Insula right **INS-R** | 3.83E-05 | 4.74E-03 | Cognition |
| Anterior cingulate gyrus right **ACG-R** | 1.00E-02 | NS | Cognition |
| Hippocampus left **HIP-L** | 9.60E-03 | NS | Cognition |
| Hippocampus right **HIP-R** | 7.61E-04 | NS | Cognition |
| Parahippocampal gyrus right **PHG-R** | 3.42E-04 | NS | Cognition |
| Lingual gyrus right **LING-R** | 1.56E-02 | 6.39E-03 | Cognition |
| Superior occipital gyrus left **SOG-L** | NS | 6.00E-03 | Cognition |
| Superior occipital gyrus right **SOG-R** | 1.59E-02 | 7.23E-03 | Cognition |
| Middle occipital gyrus left **MOG-L** | 1.02E-04 | 1.74E-02 | Cognition |
| Middle occipital gyrus right **MOG-R** | 3.49E-04 | 2.75E-04 | Cognition |
| Inferior occipital gyrus left **IOG-L** | 1.11E-02 | NS | Cognition |
| Inferior occipital gyrus right **IOG-R** | 1.25E-03 | 1.56E-03 | Cognition |
| Postcentral gyrus left **PoCG-L** | 5.43E-03 | NS | Cognition |
| Superior parietal gyrus left **SPG-L** | 8.24E-03 | 6.75E-03 | Cognition |
| Inferior parietal lobule right **IPL-R** | NS | 4.11E-03 | Cognition, Language |
| Angular gyrus left **ANG-L** | 2.80E-03 | NS | Cognition, Language |
| Caudate left **CAU-L** | 4.57E-05 | 9.18E-03 | Motor |
| Caudate right **CAU-R** | 8.69E-04 | NS | Motor |
| Putamen left **PUT-L** | 4.74E-06 | 1.66E-02 | Motor |
| Putamen right **PUT-R** | 1.84E-05 | NS | Motor |
| Pallidum left **PAL-L** | 4.17E-03 | 4.26E-03 | Motor |
| Pallidum right **PAL-R** | 1.54E-03 | NS | Motor |
| Thalamus right **THA-R** | 7.33E-04 | 2.19E-03 | Motor |
| Heschl's gyrus left **HES-L** | 5.95E-03 | NS | Language |
| Superior temporal gyrus left **STG-L** | 4.01E-04 | 5.02E-04 | Cognition, Language |
| Middle temporal gyrus left **MTG-L** | 1.16E-06 | NS | Cognition, Language |
| Middle temporal gyrus right **MTG-R** | 8.26E-04 | NS | Cognition, Language |
| Inferior temporal gyrus left **ITG-L** | 3.29E-05 | 3.93E-03 | Cognition |
| Inferior temporal gyrus right **ITG-R** | 4.01E-03 | 2.42E-03 | Cognition |

**Figure 3:**
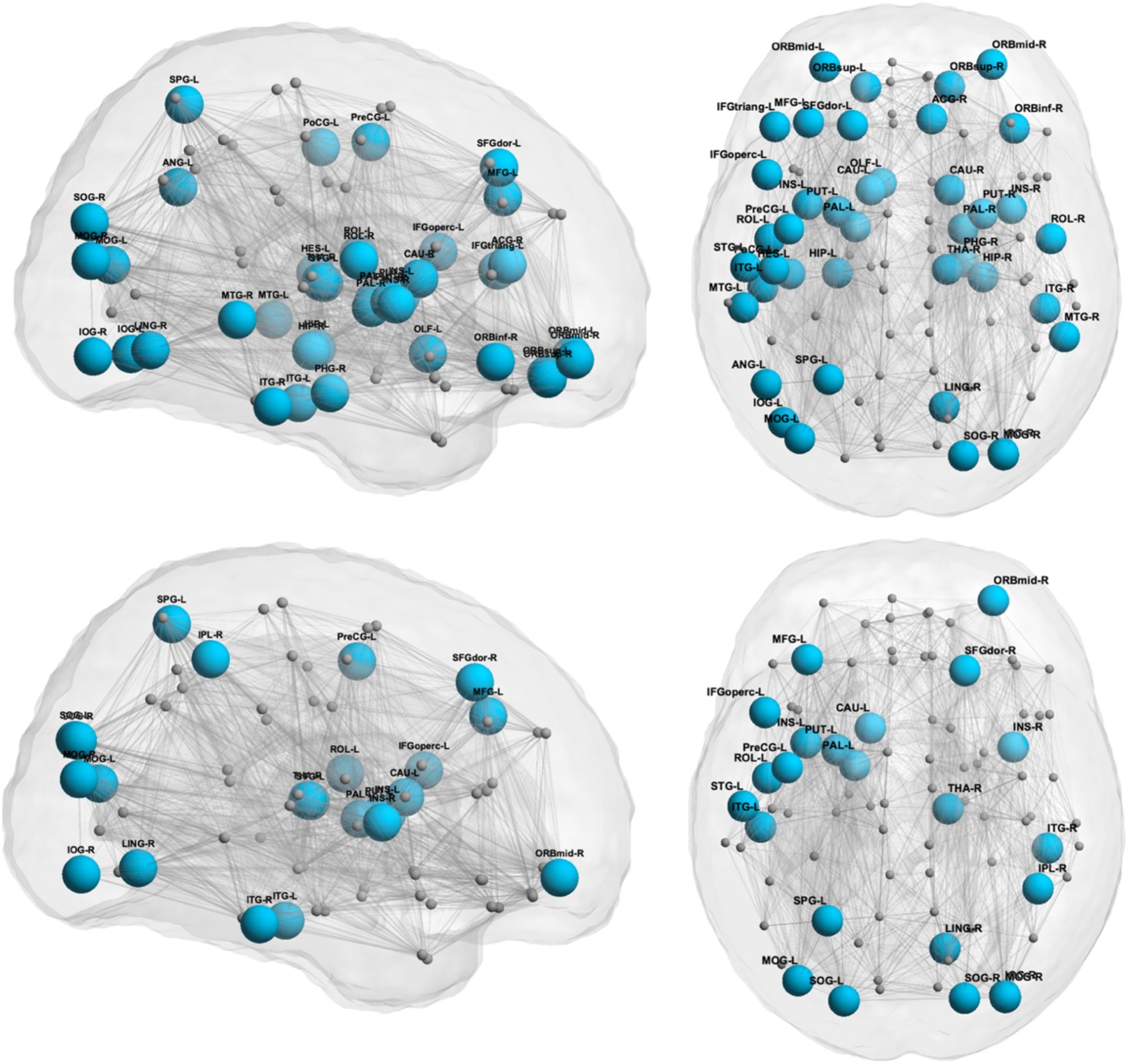
DWMA vs Regional LE and CC. Blue circles represents nodes in which objectively quantified DWMA volume was negatively correlated with local efficiency (top row) and clustering coefficient (bottom row) at term equivalent age in the very preterm brain, after correcting for postmenstrual age at MRI, gestational age, sex, structural brain abnormality score, and birth hospital and applying false discovery rate correction. The same networks are shown from a sagittal (left) and an axial (right) orientation. There were no significant positive correlations.

## Discussion

We have illustrated that in the very preterm brain, increasing volume of objectively-quantified DWMA is indicative of widespread reduced information processing ability. Increased DWMA volume correlates with decreased global efficiency, local efficiency, and clustering coefficient throughout a widely-distributed brain network that subserves cognitive, language, and motor performance. This finding supports the idea that DWMA is a pathological signal related to decreased white matter structural integrity.

Our results are consistent with the handful of studies reporting altered white matter microstructure in the presence of DWMA. In a cohort of extremely preterm infants (GA <27 weeks), Skiold and colleagues found that infants with qualitatively-diagnosed DEHSI had lower FA and higher apparent diffusion coefficient in the centrum semiovale and along the corpus collosum than infants without DEHSI ^1^. Counsell et al. reported similar results; compared to infants with normal white matter, very preterm infants with qualitatively-defined DEHSI had elevated radial diffusivity in the posterior limb of the internal capsule and the corpus collosum and elevated axial and radial diffusivity in the frontal, occipital, and periventricular white matter and the centrum semiovale ^17^. All these findings suggest that in the presence of DWMA there is either 1) reduced myelination of white matter tracts or 2) an underlying axonal abnormality involving decreased white matter integrity or disorganized white matter microstructure. Results from our postmortem histopathology case series ^19^ identified fewer oligodendroglial cells and axons in the brains of very preterm infants with DWMA, indicating that both axonal loss and reduced myelination are in fact involved in DWMA pathology. Both of these microstructural abnormalities would manifest as decreased FA, which could lead to the altered efficiency metrics seen in the current analysis.

We hypothesized that measures of information transfer ability would be reduced in networks related to cognition, language, and motor ability, given the specific impairments often seen in preterm children. Local efficiency and clustering coefficient decreased with increasing DWMA volume throughout a widely-distributed brain network, which contains many crucial hubs for cognitive, language, and motor ability (Table 2). E_loc_ and CC were reduced with increasing DWMA volume in brain regions critical to cognitive function, such as memory (e.g. the bilateral hippocampus and the right hippocampal gyrus) and the processing of complex object features (i.e. the bilateral inferior temporal gyrus). Integration metrics were also reduced in nodes crucial to language production and comprehension, including the left inferior frontal gyrus (a main hub for speech processing that contains Broca’s area) and Heschel’s gyrus (left), which contains the primary auditory cortex. Furthermore, integration measures were down-regulated with increasing DWMA in the left precentral gyrus (the location of the primary motor cortex), the left thalamus, and also bilaterally in the caudate, pallidum, and putamen, which are all parts of the brain’s motor network. See Table 2 for a complete list of the significant regions and their membership within functional networks.

Comparing our results to others in the literature raises some intriguing ideas about the antecedents of reduced efficiency in the preterm brain and also about the resilience of this network. Bouyssi-Kobar et al. used resting state functional connectivity to compare the brains of preterm and full-term infants at term-equivalent age ^27^. They recorded less integration and segregation in the preterm brain, as reflected by decreased global efficiency, local efficiency, and clustering coefficient; and they determined an association with chronic respiratory illness. We found that the same graph theory metrics are downregulated in proportion to DWMA volume, suggesting that DWMA is a robust biomarker of network disorganization in the preterm brain at term. These network alterations likely persist into early life, as Young et al. reported decreased E_glob_, E_loc_, and CC in very preterm children compared to full-term children at four years of age ^25^. On the other hand, Thompson et al. showed decreased E_glob_ but increased E_loc_ in specific brain regions in preterm children at seven years of age ^28^, which suggests that that preterm children may employ compensatory strategies to improve the efficiency of specific brain regions during early life.

Our study has a number of strengths. We recruited a regional, population-based cohort of very preterm infants, thus increasing the generalizability of our findings. This is the largest cohort ever assembled to study DWMA, which may explain why we identified such a strong association between DWMA and the global efficiency of the preterm brain (p= 8.36E-07). Unlike most prior studies, we used an semi-automated, objective method to quantify DWMA. Furthermore, by using graph theory metrics to examine higher-order brain network properties, we were able to detect significant associations with DWMA with fewer spurious results than if we had examined total white matter streamlines or mean FA between each pair of regions (There are 4005 possible regional pairs in a symmetric 90x90 network, although many are not biologically plausible). However, our study also had some limitations. Excluded subjects were more likely to have had moderate to severe brain injury, which may have introduced some sample bias. Nevertheless, our findings remained significant after controlling for structural abnormalities on MRI. We did not concurrently examine the same graph theory metrics derived from functional MRI. Such a multi-modality MRI study could corroborate our conclusions about the relationship of DWMA to network efficiency in the very preterm brain and remains a future goal of our work.

Overall, increasing volume of DWMA is correlated with reduced information processing efficiency throughout the very preterm brain, suggesting that the most common MRI finding in very preterm infants at term is associated with decreases in brain network connectivity. More work is needed to examine the relationships between DWMA, graph theory metrics, and neurodevelopmental outcomes later in life. We have begun to undertake these follow-up studies in our center.

## Data Availability

Code used in this analysis and derived data that support the conclusions of this study are available upon direct request to the corresponding author.

## Acknowledgements

This research was supported by grants R01-NS094200-05 and R01-NS096037-03 from the National Institute of Neurological Disorders and Stroke (NINDS) and R21-HD094085 from the Eunice Shriver Kennedy National Institute of Child Health and Human Development (NICHD). We sincerely thank the Cincinnati Infant Neurodevelopment Early Prediction Study (CINEPS) and the NICU fellows, nurses, and staff, and most importantly, all the study families that made this research possible.

## Author contributions

N.A.P. designed this study. V.S.P.I and N.A.P. collected the data. J.E.K., V.S.P.I., H.L., L.H., and N.A.P. analyzed the data. J.E.K. wrote the manuscript. J.E.K., V.S.P.I., H.L., L.H., and N.A.P. reviewed and edited the manuscript. All authors take full responsibility for the research contained in this article.

